# Results From the REsCue Trial: A Randomized Controlled Trial with Extended-Release Calcifediol in Symptomatic Outpatients with COVID-19

**DOI:** 10.1101/2022.01.31.22270036

**Authors:** Charles W. Bishop, Akhtar Ashfaq, Joel Z. Melnick, Enrique Vazquez-Escarpanter, Jonathan A. Fialkow, Stephen A. Strugnell, John Choe, Kamyar Kalantar-Zadeh, Noah C. Federman, David Ng, John S. Adams

## Abstract

**Importance:** The benefit of vitamin D treatment for coronavirus disease 2019 (COVID-19) remains unclear.

**Objective:** To investigate the effect of raising serum total 25-hydroxyvitamin D (25D) to 50-100 ng/mL with oral extended-release calcifediol (ERC) on time to symptom resolution in mild to moderate COVID-19.

**Design, Setting, and Participants:** A multicenter, randomized, double-blind, placebo-controlled study evaluated treatment of 160 outpatients with COVID-19 diagnosed between November 2020 and October 2021.

**Interventions:** Patients were treated for 4 weeks with ERC (30 mcg/capsule; 300 mcg on Days 1-3 and 60 mcg on Days 4-27) or placebo.

**Outcome Measures:** Primary endpoints were raising serum 25D to ≥50 ng/mL at Day 14 and resolution time for five aggregated symptoms. Secondary endpoints included resolution time for aggregated and individual symptoms as a function of serum 25D and changes in clinical biomarkers.

**Results:** 171 subjects randomized, 160 treated and 134 (65 ERC and 69 placebo) retained. Average age was 43 (range: 18-71); 59% female, 92% White, 80% Hispanic, 7% African-American, 1% Other, 76% overweight, 40% obese, 26% comorbidities, mean baseline 25D of 37±1 (SE) ng/mL. ERC increased mean 25D to 82±4 ng/mL (p<0.001) by Day 7; 88% of subjects attained a level ≥50 ng/mL; the placebo group trended lower. Resolution time for five aggregated symptoms was unchanged by ERC given that two composite non-respiratory symptoms responded poorly. Prespecified analyses showed that respiratory symptoms tended to resolve earlier when serum 25D levels reached ≥50 ng/mL, but statistical significance was limited by small sample size and non-compliance: 25D increased in seven placebo subjects (unauthorized supplementation) and none occurred in five ERC subjects (failure to dose). A post-hoc composite of three respiratory symptoms (trouble breathing, chest congestion and dry or hacking cough) resolved 3.0 days faster when 25D was elevated at Days 7 and 14 (p<0.05); chest congestion resolved 4.0 days faster with 25D increases of ≥25 ng/mL (p<0.05). Safety concerns including hypercalcemia were absent with ERC treatment.

**Conclusions and Relevance:** ERC was effective in increasing serum 25D in outpatients with COVID-19, which may have accelerated resolution of respiratory symptoms suggesting mitigation of COVID-19 pneumonia risk, findings which warrant further study.

## Introduction

Vitamin D repletion is purported to reduce the risk of infection with severe acute respiratory syndrome coronavirus 2 (SARS-CoV-2)^1,2^, mitigate severity of coronavirus disease 2019 (COVID-19)^3^ and accelerate recovery^4^. Sufficient serum total 25-hydroxyvitamin D (25D) is postulated to potentiate COVID-19 vaccine effectiveness^5^, boost innate and control adaptive immunity ^6,7^, and reduce post-infection cytokine storm^6^ and lung injury^8^. A serum 25D level of approximately 50 ng/mL is proposed as the theoretical threshold for zero mortality from COVID-19 based on regression analysis of decreasing deaths rates with rising serum 25D levels^9^.

The purported benefits of 25D repletion remain fully unsubstantiated in prospective randomized controlled trials (RCTs). In one of only a few RCTs, administration of a single oral dose of 200,000 IU of cholecalciferol (vitamin D_3_) to patients with moderate to severe COVID-19 failed to shorten hospital stays despite raising mean serum 25D from a baseline of 21.2 to 44.4 ng/mL, diminishing support for vitamin D repletion^10^.

The present RCT sought to explore the benefit of raising serum 25D to at least 50 ng/mL, a level considered by some to be of concern^11^, on time to resolution of symptoms in COVID-19 outpatients. The working hypothesis was that controlled, progressive elevation of 25D to this level with extended-release calcifediol (ERC) would safely boost the human innate immune response to SARS-CoV-2 and accelerate recovery via intracrine production of calcitriol in immune cells, such as macrophages, which co-express 25-hydroxyvitamin D-1α-hydroxylase (CYP27B1) and vitamin D receptor (VDR) in response to viral infection^12^. Apt selection of appropriate patient-reported outcomes as endpoints was hampered by the dearth of information regarding which COVID-19 symptoms respond to vitamin D treatment, leading to selection of a primary efficacy endpoint representing a “best guess” for finding a clinically meaningful vitamin D signal. To compensate, this proof-of-concept study explored a broad range of symptoms and used numerous prespecified and post-hoc analyses to characterize potential signals.

## METHODS

This multicenter trial (NCT04551911) titled “A Randomized, Double-Blind Placebo-Controlled Study to Evaluate the Safety and Efficacy of **R**ayaldee (calcifediol) **E**xtended-relea**s**e **C**aps**u**l**e**s to Treat Symptomatic Patients Infected with SARS-CoV-2 (**REsCue**)” enrolled symptomatic COVID-19 outpatients from 10 sites across the United States and randomized them in a 1:1 ratio for 4 weeks of treatment with ERC (30 mcg/capsule) or placebo and a 2-week follow-up. ERC was chosen as the test intervention because it has been previously shown to safely and effectively raise serum 25D to targeted levels of 50-100 ng/mL and suppress elevated intact parathyroid hormone (iPTH) when administered at 30 mcg/day and escalating, as needed, to 60 mcg/day in patients with stage 3 or 4 chronic kidney disease (CKD).^13^ Dosing with ERC in the current study was designed to raise 25D more quickly, but in a controlled, progressive manner, to this same target range well before Day 7, beginning with 300 mcg (10 capsules) on each of Days 1, 2 and 3 followed by 60 mcg (2 capsules) on Days 4 through 27.Doses were administered at bedtime after fasting for three hours following dinner, and subjects were instructed to remain fasting for 3 hours after dosing. Thirty-four COVID-19 symptoms were self-reported daily using the FLU-PRO Plus^©^ questionnaire, an outcome tool validated for respiratory tract viral infections^14^. Symptoms were scored using multipoint scales ranging, most frequently, from 0 to 4. Blood samples and safety assessments were obtained at baseline and 7-day intervals.

### Participants

All patients provided written informed consent before participation in the study. Patients were enrolled between November 2, 2020 and August 27, 2021 after testing positive for SARS-CoV-2 infection within the previous three days via reverse transcription polymerase chain reaction (RT-PCR) or another substitutable FDA-authorized test. They were ≥18 years of age, and had mild to moderate COVID-19 (absence of clinical signs indicative of more severe disease such as oxygen saturation <94% or respiration rate >30 bpm). They were required to have symptoms during screening with mean scores of ≥1.5 for each of the chest/respiratory and body/systemic domains of the FLU-PRO Plus^©^ questionnaire. They were instructed to forgo use of vitamin D supplements during the study. Patients were excluded if: they were pregnant or breastfeeding; had recently taken systemic glucocorticoid medications; had a recent history of primary hyperparathyroidism, kidney stones, hypercalciuria or hypercalcemia, cardiovascular disease, poorly controlled hypertension, arrhythmias, chronic granuloma-forming disease or chronic liver disease; history in the past five years of multiple myeloma or carcinoma of the breast, lung or prostate; any surgical or medical condition that might significantly alter the absorption, distribution, metabolism, or excretion of vitamin D; ongoing treatment with thiazide diuretics; history of hyperphosphatemia, hyperuricemia or gout; estimated glomerular filtration rate (eGFR) < 15 mL/min/1.73m^2^; or serum calcium ≥ 9.8 mg/dL within the last three months.

### Independent Oversight and Blinding

The study was approved by Advarra (Columbia, MD), a central Institutional Review Board (IRB), and overseen by an independent five-member Data Safety Monitoring Board (DSMB) comprised of an expert in viral disease pathogenesis, a nephrologist, a biostatistician, a pulmonary medicine expert and an endocrinologist experienced in clinical safety monitoring. The monitor reduced the maintenance dose to one capsule per day at Day 21 if serum total 25D exceeded 100 ng/mL or serum total calcium (corrected for low albumin) exceeded 10.5 mg/dL. Subjects, study personnel and the sponsor (and its designees, with the exception of the DSMB and three unblinded members of the data management team) were blinded until after database lock to treatment assignments and to serum vitamin D metabolite laboratory data.

### Outcome Measures

One primary endpoint was attainment of the targeted serum total 25D level by Day 14. A second was time to resolution of five composite COVID-19 symptoms (trouble breathing, chest congestion, dry or hacking cough, body aches or pains, chills or shivering). These symptoms were part of the chest/respiratory and body/systemic domains of the FLU-PRO Plus^©^ questionnaire for which mean scores of ≥1.5 were required for enrollment. The composite respiratory symptoms (trouble breathing, chest congestion, dry or hacking cough) were considered indicative of pulmonary compromise. The other two composite symptoms (body aches or pains, chills or shivering) were considered indicative of systemic inflammatory responses to viral infection. An aggregate score for the primary endpoint related to five COVID-19 symptoms was selected in view of the observed heterogeneity in COVID-19 symptom profiles; not all subjects experienced the same symptoms or degree of symptom severity. A score of ≤5, or a mean of ≤1 for each composite symptom, was deemed appropriate because the threshold of ≤1 described minor symptom severity (at worst “a little bit”) which seemed unlikely to presage a poor outcome. Resolution was defined as a reduction in the total baseline score (maximum of 20) for the five symptoms to or below 5 for a minimum of three consecutive days. Secondary endpoints included time to resolution of each composite symptom and of aggregated symptoms as a function of serum 25D.

Safety endpoints included adverse events detected by physical examinations, vital signs, electrocardiograms, hematology and clinical chemistries. Special attention was given to changes in serum calcium and phosphorus, and eGFR, which presage potential hypercalcemia, hyperphosphatemia and kidney damage. Exploratory endpoints included changes in serum 1,25-dihydroxyvitamin D (1,25D) and LL37 cathelicidin antimicrobial peptide, and plasma iPTH.

### Laboratory Procedures

BioReference Laboratories (Elmwood Park, NJ) analyzed serum total 25D by LC-MS, plasma iPTH by electrochemiluminescence (Roche Elecsys) and serum total 1,25D by chemiluminescence (DiaSorin Liason). Syneos Health (Quebec, Canada) analyzed serum LL37 by ELISA (LSBio). The circulating neutrophil:lymphocyte ratio was obtained from the complete blood cell count as routinely performed.

### Statistical Analysis

Results from analysis of the per-protocol (PP) population rather than the full analysis set (FAS) are presented herein as the study merely sought to identify a clinically meaningful vitamin D signal in the treatment of COVID-19 outpatients and guide the design of a larger, confirmatory follow-on study. The two primary endpoints were tested hierarchically to maintain an overall one-sided alpha level of 0.025. The first, attainment of serum 25D levels of ≥50 ng/mL, was assessed with a Chi-square statistic. The second, number of days to resolution of five aggregated symptoms, was analyzed with a Cox proportional hazards model with three covariates considered to have possible influence on treatment efficacy: baseline FLU-PRO Plus^©^ score for the five aggregated symptoms, baseline serum 25D, and body weight. The log-rank test was used post-hoc to compare Kaplan-Meier curves examining time to resolution of individual or aggregated symptoms as a function of increase (versus lack thereof) in serum total 25D at both Days 7 and 14.Subjects who did not achieve symptom resolution prior to ending participation in the study (3 to 19% depending on the symptom being evaluated; mean = 8.8%) were right censored after the last day when data were obtained. Other efficacy endpoints were analyzed at each time point using appropriate *t*-tests.

## RESULTS

### Patients

A total of 241 patients provided written informed consent and were screened for eligibility (**Figure 1**). Of these, 171 met all selection criteria and were randomized to one of the two treatment groups. The most common reason for failing screening was insufficient severity of COVID-19 symptoms. Five subjects withdrew consent prior to dosing, and six failed to receive the shipment of study drug, leaving 160 subjects (80 per treatment group) who received at least one dose of study drug (safety population). Eight subjects assigned to ERC and 5 subjects assigned to placebo achieved symptom resolution prior to dosing and were excluded from analysis, leaving 147 subjects (72 ERC and 75 placebo) in the FAS. Thirteen subjects had major deviations from the protocol (e.g., less than 80% dosing compliance as documented in daily diaries) prior to resolution of symptoms and were excluded from the PP population, which consisted of 134 subjects (65 ERC and 69 placebo).

**Figure 1:**
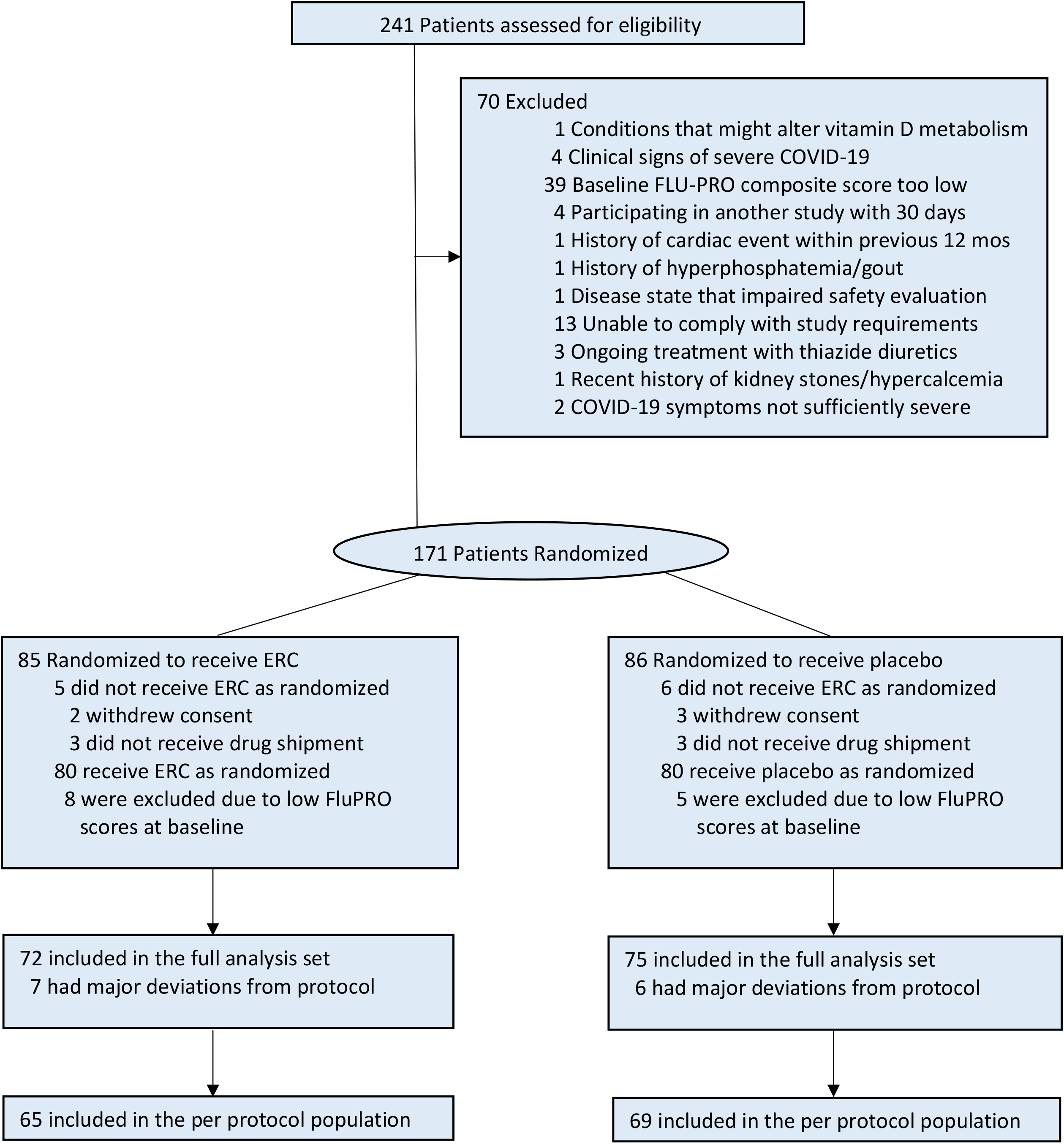
Consort Diagram.

The average age of PP subjects was 43 (range: 18-71); 59% were female, 92% White, 7% African-American, 1% Other, 76% were overweight and 40% obese based on body mass index (BMI) greater than 25 and 30, respectively. Approximately 26% had comorbidities, most commonly hypertension. Baseline mean serum total 25D was 37±1 (SE) ng/mL, with 95% or 72% of subjects having values of at least 20 or 30 ng/mL, respectively, two thresholds widely viewed as lower limits of adequacy^11,15^. Baseline characteristics of the PP population are shown in **Table 1;** no significant differences were evident between treatment groups.

**Table 1:**
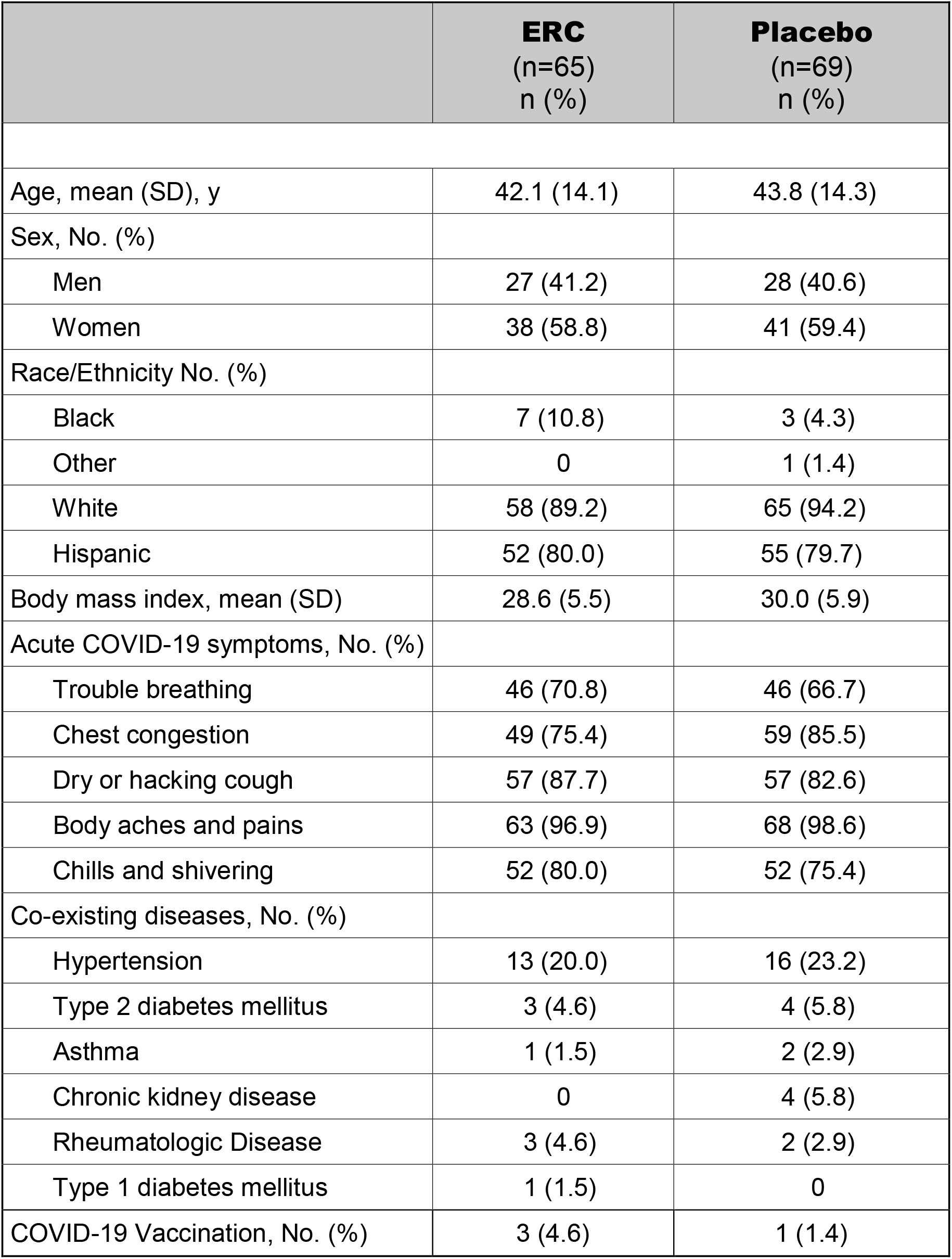
Baseline Characteristics of Per-protocol Subjects.

|  | **ERC**<br>(n=65)<br>n (%) | **Placebo**<br>(n=69)<br>n (%) |
| --- | --- | --- |
| Age, mean (SD), y | 42.1 (14.1) | 43.8 (14.3) |
| Sex, No. (%) |  |  |
| Men | 27 (41.2) | 28 (40.6) |
| Women | 38 (58.8) | 41 (59.4) |
| Race/Ethnicity No. (%) |  |  |
| Black | 7 (10.8) | 3 (4.3) |
| Other | 0 | 1 (1.4) |
| White | 58 (89.2) | 65 (94.2) |
| Hispanic | 52 (80.0) | 55 (79.7) |
| Body mass index, mean (SD) | 28.6 (5.5) | 30.0 (5.9) |
| Acute COVID-19 symptoms, No. (%) |  |  |
| Trouble breathing | 46 (70.8) | 46 (66.7) |
| Chest congestion | 49 (75.4) | 59 (85.5) |
| Dry or hacking cough | 57 (87.7) | 57 (82.6) |
| Body aches and pains | 63 (96.9) | 68 (98.6) |
| Chills and shivering | 52 (80.0) | 52 (75.4) |
| Co-existing diseases, No. (%) |  |  |
| Hypertension | 13 (20.0) | 16 (23.2) |
| Type 2 diabetes mellitus | 3 (4.6) | 4 (5.8) |
| Asthma | 1 (1.5) | 2 (2.9) |
| Chronic kidney disease | 0 | 4 (5.8) |
| Rheumatologic Disease | 3 (4.6) | 2 (2.9) |
| Type 1 diabetes mellitus | 1 (1.5) | 0 |
| COVID-19 Vaccination, No. (%) | 3 (4.6) | 1 (1.4) |

### Outcomes

Mean serum 25D levels increased with ERC treatment to 82±4 (SE) ng/mL (p<0.001) by Day 7 and remained elevated for the duration of the study, with 88% of subjects attaining the targeted level of ≥50 ng/mL (**Figure 2A**). Attainment of this endpoint was unaffected by body weight or BMI. In contrast, mean serum 25D trended lower with placebo treatment. Ten subjects receiving ERC required dose reductions at Day 21 due to 25D levels rising above 100 ng/mL. ERC treatment produced similar mean increases in serum 25D in each baseline 25D category (**Figure 2B**).

**Figure 2:**
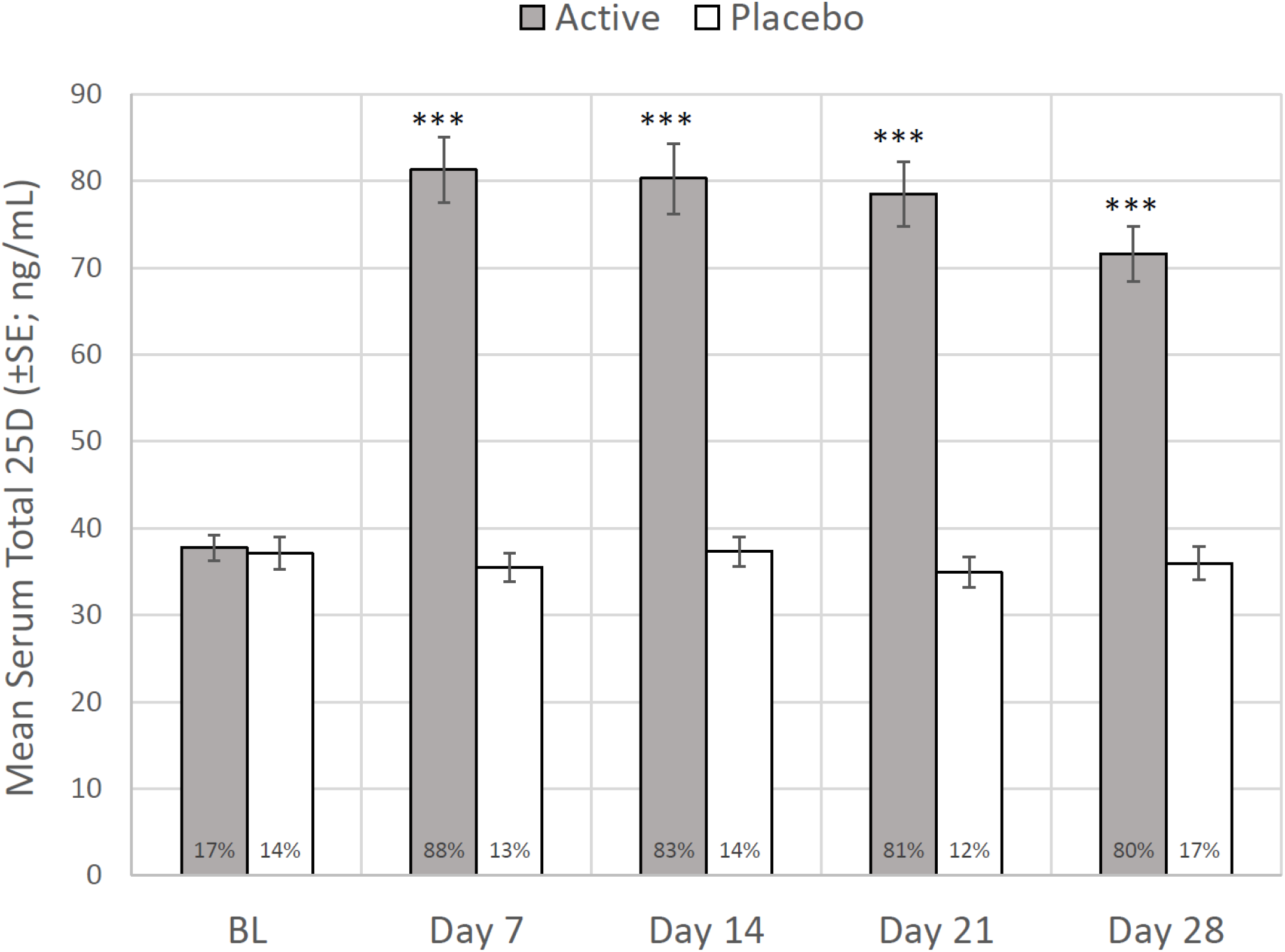
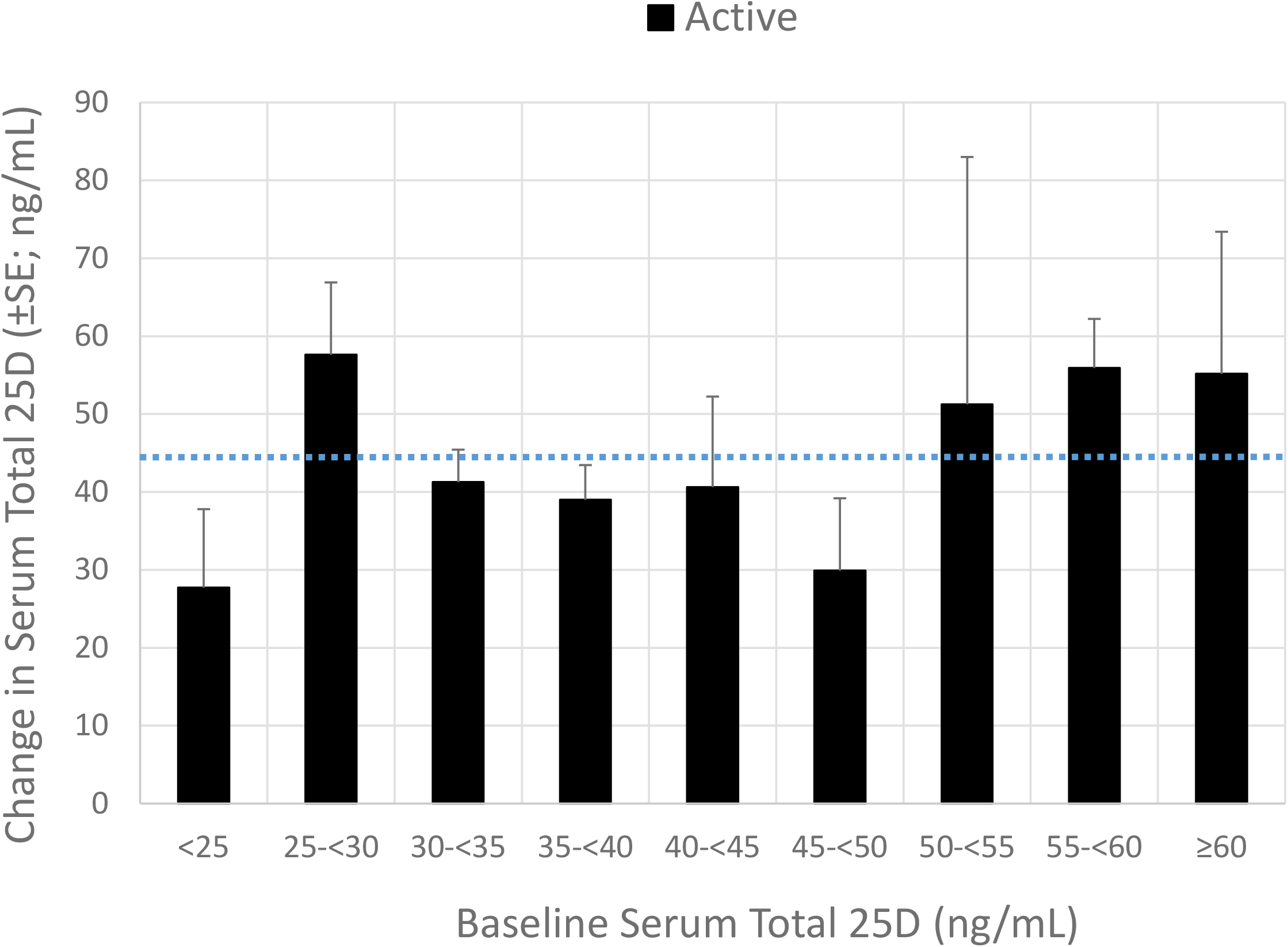
**(Panel A): Mean (SE) Serum Total 25-hydroxyvitamin D by Study Day and Treatment Group (Per-protocol Population)**. Percentages at the base of each bar indicate the proportion of subjects achieving serum 25D levels of at least 50 ng/mL. Asterisks indicate significant differences between treatment groups (p<0.001). **(Panel B): Mean (SE) Increases in Serum Total 25-hydroxyvitamin D with ERC Treatment by Study Day (Per-protocol Population)**. The horizontal dotted line indicates the mean increase for all treated subjects. * * *Significantly different from Placebo, p < 0.001

The time to resolution for the five aggregated COVID-19 symptoms was statistically unchanged by ERC given that two composite non-respiratory symptoms responded poorly (**Figure 3A**). Prespecified analyses showed that the other three respiratory symptoms tended to resolve sooner, for example, chest congestion (**Figure 3B**), when serum 25D levels reached at least 50 ng/mL; however, significance of the changes was limited by small sample size and dosing non-adherence: 25D increases occurred in seven placebo subjects (unauthorized supplementation) and none occurred in five ERC subjects (failure to dose). These three respiratory symptoms (trouble breathing, chest congestion and dry or hacking cough), when analyzed together post-hoc, resolved 3.0 days faster (**Figure 3C**) on average when serum 25D was elevated above baseline at Days 7 and 14 (p<0.05); chest congestion resolved 3.4 (**Figure 3D**) days faster (p<0.05). Resolution time for chest congestion was accelerated by 4.0 days with 25D increases of at least 25 ng/mL (p<0.05).

**Figure 3:**
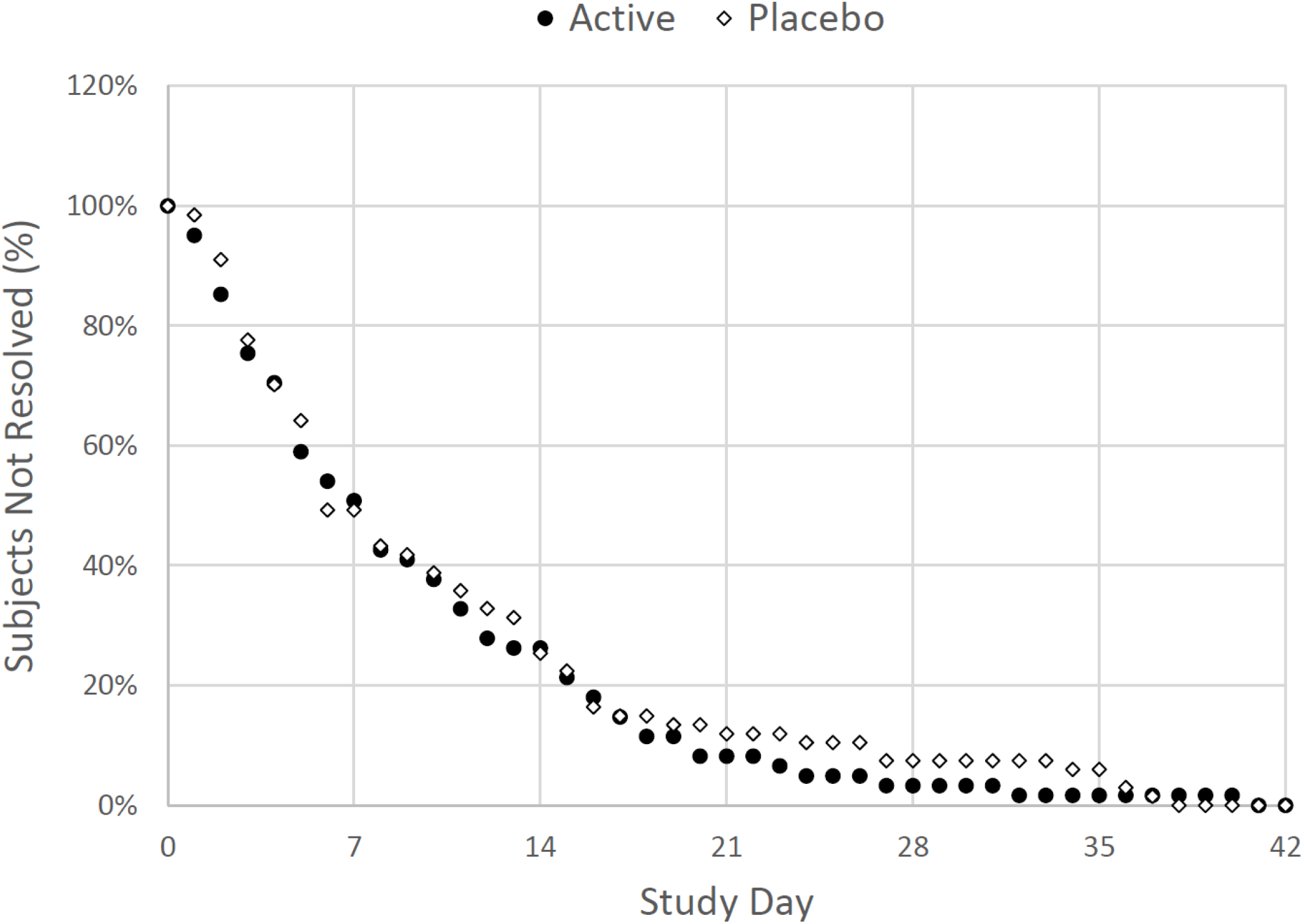
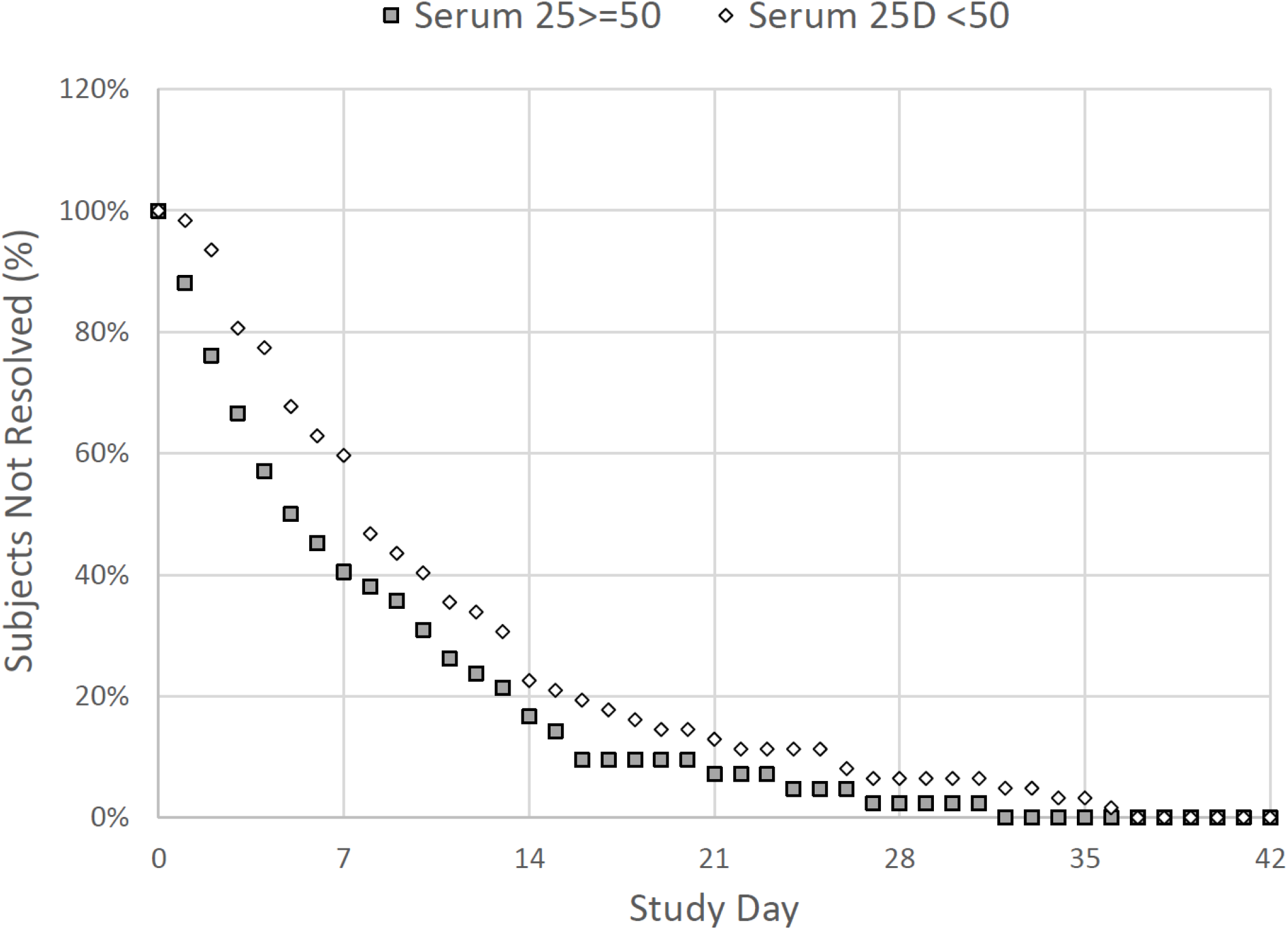
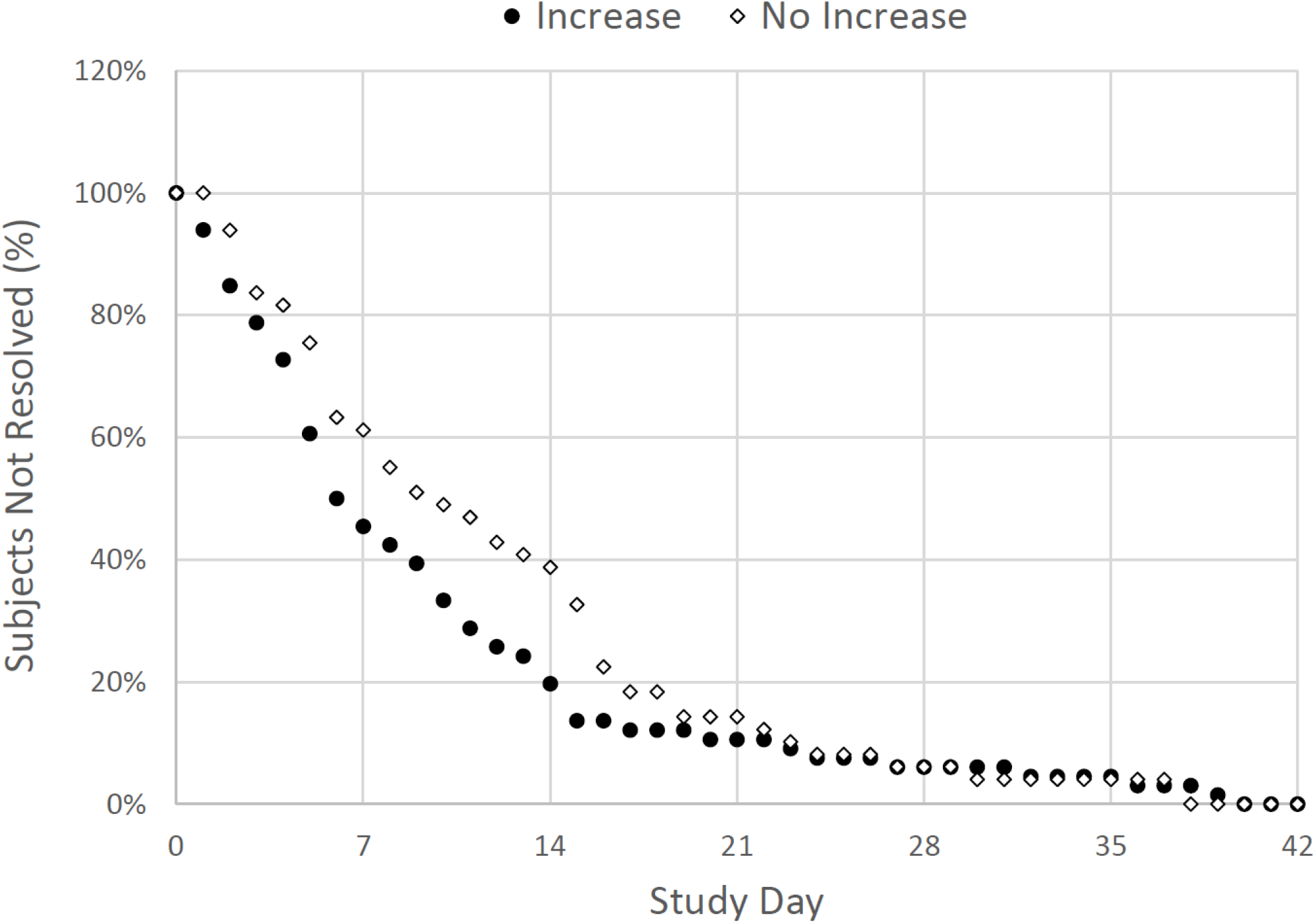
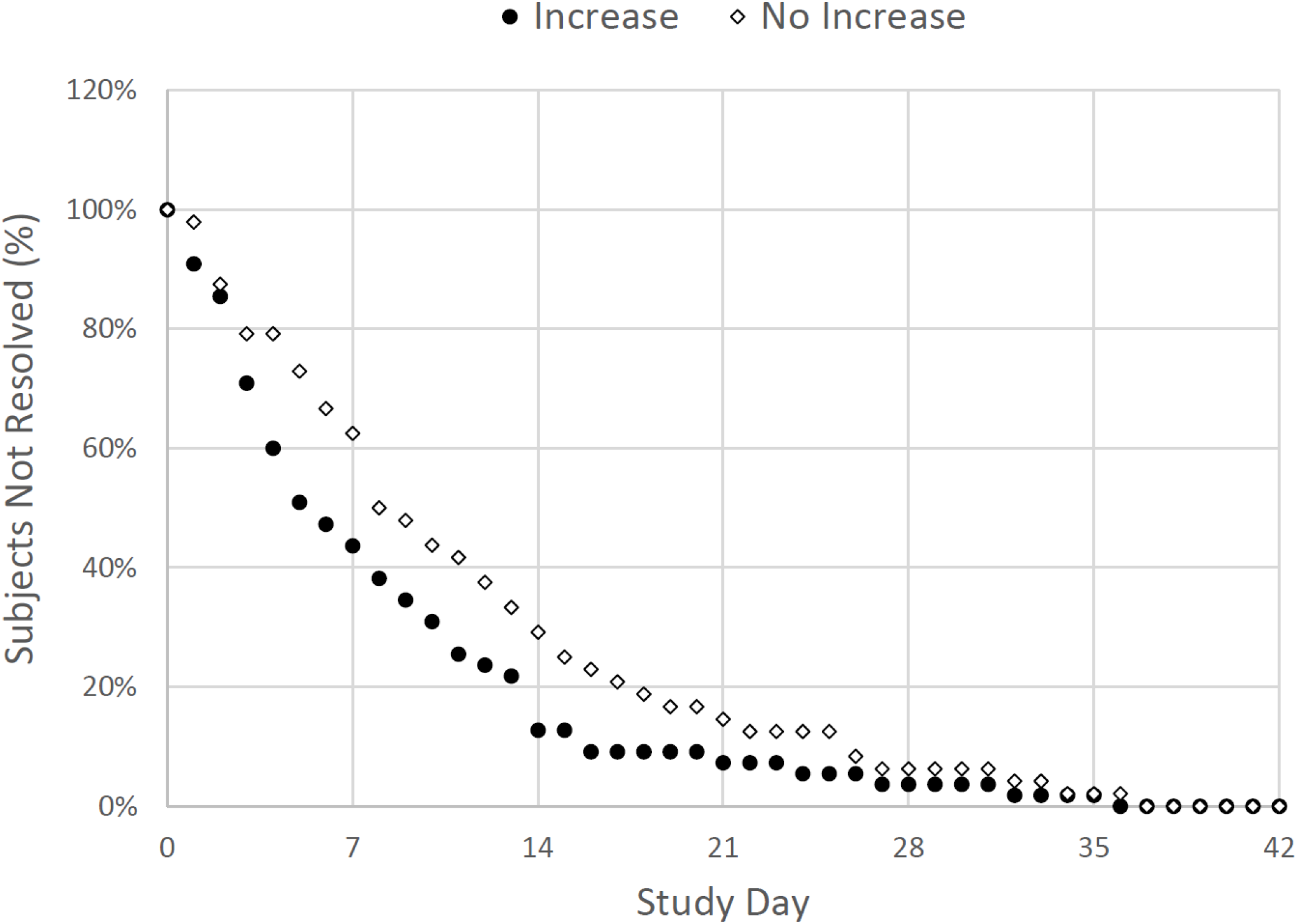
**(Panel A): Kaplan-Meier Curves Displaying the Time to Resolution of Five Aggregated Symptoms (Trouble Breathing, Chest Congestion, Dry or Hacking Cough, Body Aches or Pains, Chills or Shivering) by Treatment Group (Per-protocol Population)**. Active group: symptoms resolved in 61 subjects but not in 4 who were right-censored. Placebo group: symptoms resolved in 67 subjects but not in 2 who were right-censored. The difference between the plotted curves is not statistically significant. **(Panel B): Kaplan-Meier Curves Displaying the Time to Resolution of Chest Congestion in Subjects Achieving at both Days 7 and 14 Serum Total 25-Hydroxyvitamin D Levels of At Least 50 ng/mL Versus Less than 50 ng/mL (Per-protocol Population)**. High 25D group: symptoms resolved in 42 subjects but not in 8 who were right-censored. Low 25D group: symptoms resolved in 62 subjects but not in 3 who were right-censored. The difference between the plotted curves is not statistically significant. A total of 19 subjects did not have chest congestion at treatment initiation (Day 1) and were excluded from analysis. **(Panel C): Kaplan-Meier Curves Displaying the Time to Resolution of a Composite of Three Respiratory Symptoms (Trouble Breathing, Chest Congestion, Dry or Hacking Cough) in Subjects Achieving at both Days 7 and 14 Increases Versus No Increases in Serum Total 25-** **Hydroxyvitamin D Levels (Per-protocol Population)**. Increase group: symptoms resolved in 66 subjects but not in 4 who were right-censored. No increase group: symptoms resolved in 49 subjects but not in 5 who were right-censored. The difference between the plotted curves is statistically significant (P<0.05). A total of 10 subjects did not have a total score of >3 at treatment in at treatment initiation (Day 1) and were excluded from analysis. **(Panel D): Kaplan-Meier Curves Displaying the Time to Resolution of Chest Congestion in Subjects Achieving at both Days 7 and 14 Increases Versus No Increases in Serum Total 25-Hydroxyvitamin D Levels (Per-protocol Population)**. Increase group: symptoms resolved in 56 subjects but not in 7 who were right-censored. No increase group: symptoms resolved in 48 subjects but not in 4 who were right-censored. The difference between the plotted curves is statistically significant (P<0.05). A total of 19 subjects did not have chest congestion at treatment initiation (Day 1) and were excluded from analysis.

The sample size of the study was too small to evaluate the effect of treatment on incidence of urgent care visits, oxygen saturation below 94% and hospitalizations. Changes in laboratory measurements during treatment are summarized in **Table 2**. Serum total 1,25D, serum LL37 and the neutrophil:lymphocyte ratio all trended upward with ERC treatment and downward with placebo, while serum calcium and phosphorus, plasma iPTH and eGFR remained stable. Mean serum calcium in the ERC group was unaffected by baseline or post-treatment serum 25D level. Episodes of hypercalcemia or hyperphosphatemia were not observed, even in subjects whose serum 25D exceeded 100 ng/mL. Other safety endpoints showed no clinically meaningful changes with treatment.

**Table 2:** Changes in Clinical Chemistries From Baseline to Day 7 and Day 14 by Treatment Group.

| Lab test<br>Mean (SD) | Treatment | Baseline<br>(Visit 1) | Day 7<br>(Visit 2) | Day 14<br>(Visit 3) |
| --- | --- | --- | --- | --- |
| 1,25-Dihydroxyvitamin D (pg/mL) | CTAP101 | 71.1 (29.6) | 78.4 (34.7) * | 74.8 (23.8)** |
|  | Placebo | 74.1 (25.0) | 66.6 (20.5) | 59.4 (21.7) |
| 25-Hydroxyvitamin D (ng/mL) | CTAP101 | 37.7 (12.1) | 81.8 (30.5)** | 79.9 (32.6)** |
|  | Placebo | 37.1 (15.6) | 34.8 (13.8) | 36.4 (13.5) |
| Calcium Corrected (mg/dL) | CTAP101 | 8.81 (0.37) | 9.03 (0.38) | 8.99 (0.34) |
|  | Placebo | 8.74 (0.37) | 8.90 (0.33) | 9.01 (0.35) |
| Glomerular Filtration Rate (mL/min/1.73m<sup>2</sup>) | CTAP101 | 98.0 (17.07) | 99.1 (16.4) | 98.2 (14.4) |
|  | Placebo | 99.2 (19.4) | 100.5 (19.9) | 101.3 (18.6) |
| LL37 (ng/mL) | CTAP101 | 1.61 (1.41) | 1.67 (1.43) | 1.76 (1.68) |
|  | Placebo | 1.66 (1.63) | 1.69 (1.43) | 1.63 (1.32) |
| Neutrophil/Lymphocyte ratio (%) | CTAP101 | 2.01 (1.24) | 2.12 (1.05) | 2.05 (1.00) |
|  | Placebo | 2.04 (1.07) | 2.00 (1.06) | 2.02 (0.73) |
| Parathyroid Hormone, Intact (pg/mL) | CTAP101 | 28.8 (14.7) | 27.6 (15.9) | 26.6 (11.5) |
|  | Placebo | 30.2 (13.0) | 33.8 (15.1) | 33.4 (14.6) |
| Phosphorus (mg/dL) | CTAP101 | 3.36 (0.49) | 3.43 (0.61) | 3.44 (0.54) |
|  | Placebo | 3.37 (0.65) | 3.28 (0.52) | 3.41 (0.61) |

*Differences between treatment groups are significant: \* $P < 0.05$ or \*\* $P < 0.001$*

## DISCUSSION

This randomized, double-blind, placebo-controlled clinical trial showed that ERC treatment was effective in increasing serum total 25D to levels of at least 50 ng/mL, which may have yielded significantly shorter resolution times for three aggregated respiratory symptoms (trouble breathing, chest congestion and dry or hacking cough) commonly observed in patients with mild to moderate COVID-19. Of these symptoms, chest congestion showed the greatest response, resolving 3.4 to 4.0 days earlier, depending on the degree to which 25D was elevated. These findings suggest that raising serum 25D with ERC may mitigate, without adverse effects, the risk of COVID-19 pneumonia.

A possible mechanism^12^ underlying these findings is induction of endogenous human antimicrobial peptides, such as LL37, that can boost the host’s immune response to the virus. Serum LL37, which trended upwards in this study, is the product of the cathelicidin antimicrobial peptide (*CAMP*) gene and is secreted from monocyte/macrophages, dendritic cells and neutrophils in response to viral or bacterial infection. The innate immune response is initiated and perpetuated in antigen-presenting cells by pathogen-associated molecular patterns derived from the viral capsid and other unique signatures of the infecting virus interacting with pattern recognition receptors (e.g., Toll-like receptors [TLR]). TLR activation leads to up-regulation of expression of the intracellular CYP27B1 and VDR; the former event up-regulates the enzymatic conversion of the prohormone calcifediol to its active metabolite, calcitriol, which can then engage the VDR in an intracrine mode and control *CAMP* gene expression. Only in higher primates, including man, is the *CAMP* gene regulated by calcitriol^17^.Serum 25D of approximately 50 ng/mL or more is thought sufficient to support intracellular generation of 1,25D, activation of the VDR, transactivation of the *CAMP* gene and production and release of LL37 to combat SARS-CoV-2 proliferation in the host^16^. Serum levels of 1,25D and LL37 were observed to trend upward with ERC treatment, reflecting their elevated intracellular concentrations in the inflammatory microenvironment of the lung. The circulating neutrophil:lymphocyte ratio, a purported biomarker of disease activity in COVID-19^18^, went up in this study, but declined significantly with immediate-release calcifediol (IRC) in a previously reported RCT^19^ conducted in patients hospitalized with more severe COVID-19. Aside from this biomarker, no other significant differences between IRC and placebo treatment were reported in that earlier RCT. Possible explanations for these divergent findings include fewer participants (28 active, 34 placebo), more severe COVID-19, more rapid and proximal release of calcifediol, and lower achieved mean (SD) serum 25D level (42.0±13.7 ng/mL). Release of calcifediol from ERC is gradual, continues over a period of 12 hours (based on *in vitro* dissolution), and likely occurs primarily in the colon.

The current study differed from previous evaluations of vitamin D supplementation in COVID-19 patients in two ways. First, it targeted achievement of a high serum 25D exposure (50-100 ng/mL) with downward dose adjustment, as needed. This approach was chosen in view of the reported inverse relationship between baseline 25D and observed increase with vitamin D supplementation^20^ (a relationship that was not observed in this study), and the known adverse impact of adipose tissue on 25D bioavailability that can be readily overcome with ERC, but not with vitamin D supplements^21^. Prescription and non-prescription supplements of cholecalciferol (vitamin D_3_) and ergocalciferol (vitamin D_2_) are fat-soluble, poorly absorbed in the intestine, accumulate preferentially in adipose tissue^22,23^, possess low affinities for the serum vitamin D binding protein (DBP)^22^ and are poorly mobilized from adipose into circulation for hepatic activation^24^. Such supplements have multi-day delays in raising serum 25D^25^, and prove to be unreliable in raising serum 25D in overweight or obese patients who are at high risk for COVID-19^15,26^. Further, hepatic 25-hydroxylase activity is reduced in obesity, also blunting the intended elevation of serum 25D^27^. In contrast, calcifediol requires no hepatic activation^28^, is more water soluble^29^ and avidly binds to DBP^30^, reducing accumulation in adipose tissue^31^ and enabling ready availability to peripheral tissues, including virus-activated immune cells containing CYP27B1. Second, consistent with the excellent safety profile established for ERC in patients with stage 3-4 CKD^13^, the slow release formulation of ERC safely achieved high serum 25D exposures (50 to 100 ng/mL) that have been considered concerning for vitamin D supplements^11^.

### Strengths

The strengths of this study are the randomized, double-blind, placebo-controlled design; targeting, safely achieving and maintaining uncommonly high and sustained serum 25D levels of ≥50 ng/mL; examination of a broad spectrum of symptoms commonly associated with COVID-19; and in-depth monitoring of patient outcomes and safety via weekly in-person clinic visits.

### Limitations

The inability of this study to show more significant differences in time to resolution for respiratory symptoms is likely due to inadequate power (small sample size); the modest severity of COVID-19 in the enrolled subjects; the fact that 17% of subjects assigned to placebo treatment had, unexpectedly, serum 25D levels above 50 ng/mL at baseline; dosing noncompliance (four previous but unpublished pharmacokinetic studies of a single 900 mcg ERC loading dose in healthy volunteers have shown that no subject failed to achieve an increase in serum 25D of at least 6 ng/mL); and, varying periods of time between onset of symptoms and diagnosis in of COVID-19 as well as between diagnosis and initiation of ERC treatment. The observed positive effect of 25D elevation on resolution of COVID-19 respiratory symptoms is based on a post-hoc analysis and needs to be confirmed in a larger study with an appropriate prespecified endpoint.

## CONCLUSIONS

ERC was effective in increasing serum total 25D to levels of at least 50 ng/mL in outpatients with COVID-19 that may have accelerated resolution of respiratory symptoms, suggesting mitigation of COVID-19 pneumonia risk. The positive findings from this RCT warrant confirmation in additional larger studies.

## Data Availability

All top-line data produced in the present study are contained in the manuscript. Additional analyses are ongoing as further laboratory data become available.

## Article Information

## Author Contributions

Drs. Bishop, Ashfaq, Melnick, Ng and Choe had full access to data that are currently available from the study and take responsibility for the integrity of the data and the accuracy of the data analysis.

*Concept and design*: Drs. Bishop, Ashfaq, Melnick, Federman and Adams.

*Acquisition, analysis and interpretation*: All authors.

*Drafting of the manuscript*: All authors.

*Critical revision of the manuscript for important intellectual content*: All authors.

*Statistical analysis*: Drs. Bishop, Strugnell, Choe and Ng.

*Supervision*: Drs. Ashfaq and Melnick.

## Data Sharing Statement

All top-line data produced in the present study are summarized in the manuscript. Additional analyses are ongoing as further laboratory data become available.

## REFERENCES

1. Kaufman HW, Niles JK, Kroll MH et al. SARS-CoV-2 positivity rates associated with circulating 25-hydroxyvitamin D levels. Plos One 2020;15(9):e0239252. doi: 10.1371/journal.pone.0239252.

2. Grant WB, Lahore H, McDonnell SL, et al. Evidence that vitamin D supplementation could reduce risk of Influenza and COVID-19 infections and deaths. Nutrients 2020;12(4):988. doi: 10.3390/nu12040988.

3. Jude EB, Ling SF, Allcock R, et al. Vitamin D deficiency is associated with higher hospitalization risk from COVID-19: a retrospective case-control study. J Clin Endocrinol Metab 2021;106(11):e4708–e4715. doi: 10.1210/clinem/dgab439.

4. Wang Z, Joshi A, Leopold K, et al. Association of vitamin D deficiency with COVID-19 infection severity: systematic review and meta-analysis. Clin Encrinol 2021;10.1111/cen.14540. doi: 10.1111/cen.14540.

5. Chiu S-K, Tsai K-W, Wu C-C, et al. Putative role of vitamin D for COVID-19 vaccination. Int J Mol Sci 2021; 22(16):8988. doi: 10.3390/ijms22168988.

6. Bilezikian JP, Bikle D, Hewison M, et al. Mechanisms in endocrinology:vitamin D and COVID-19. Eur J Endocrinol 2020;183(5):R133–R147. doi: 10.1530/EJE-20-0665.

7. Martens P-J, Gysemans C, Verstuyf A et al. Vitamin D’s effect on immune function. Nutrients 2020;12(5):1248. doi: 10.3390/nu12051248.

8. Ghelani D, Alesi S, Mousa A. Vitamin D and COVID-19: an overview of recent evidence. Int J Mol Sci 2021;22(19):10559. doi: 10.3390/ijms221910559.

9. Borsche L, Glauner B, von Mendel J. COVID-19 mortality risk correlates inversely with vitamin D_3_ status, and a mortality rate close to zero could theoretically be achieved at 50 ng/mL 25(OH)D_3_: results of a systematic review and meta-analysis. Nutrients 2021;13:3596.doi: 10.3390/nu13103596.

10. Murai IH, Fernandes AL, Sales LP, et al. Effect of a single high dose of vitamin D_3_ on hospital length of stay in patients with moderate to severe COVID-19: a randomized clinical trial. JAMA 2021;doi: 10.3390/nu13103596

11. Institute of Medicine. 2011. Dietary reference intakes for calcium and vitamin D. Washington, DC: the National Academies Press. doi:10.17226/13050.

12. White JH. Emerging roles of vitamin D-induced antimicrobial peptides in antiviral innate immunity. Nutrients 2022;14:284. doi:10.3390/nu14020284.

13. Sprague SM, Crawford PW, Melnick JZ et al. Use of extended-release calcifediol to treat secondary hyperparathyroidism in stages 3 and 4 chronic kidney disease. Am J Nephrol 2016;44:316–325.

14. Powers JH, Guerrero ML, Leidy NK, et al. Development of the Flu-PRO: a patient-reported outcome (PRO) instrument to evaluate symptoms of influenza. BMC Infect Dis 2016;16:1. doi: 10.1186/s12879-015-1330-0.

15. Holick MF, Binkley NC, Bischoff-Ferrari HA, et al. Evaluation, treatment, and prevention of vitamin D deficiency: an Endocrine Society clinical practice guideline. J Clin Endocrinol Metab 2011;96:1911–30.

16. Grant WB. Vitamin D’s role in reducing risk of SARS-CoV-2 and COVID-19 incidence, severity, and death. Nutrients 2022;14:183. doi:10.3390/nu14010183.

17. Gombart, AF, Saito T, Koeffler HP. Exaptation of an ancient Alu short interspersed element provides a highly conserved vitamin D-mediated innate immune response in humans and primates. BMC Genomics 2009;10:321. doi:10.1186/1471-2164-10-321.

18. Qin C, Zhou L, Hu Z, et al. Dysregulation of immune response in patients with coronavirus 2019 (COVID-19) in Wuhan, China. Clin Infect Dis 2020;71:762–768.

19. Maghbooli Z, Sahraian MA, Jamalimoghadamsiahkali S, et al. Treatment with 25-hydroxyvitamin D_3_ (calcifediol) is associated with a reduction in the blood neutrophil-to-lymphocyte ratio marker of disease severity in hospitalized patients with COVID-19: a pilot multicenter, randomized, placebo-controlled, double-blinded clinical trial. Endocr Pract 2021;27:1242–1251.

20. Didriksen A, Grimnes G, Hutchinson MS, et al. The serum 25-hydroxyvitamin D response to vitamin D supplementation is related to genetic factors, BMI, and baseline levels. Eu J Endocrinol 2013;169:559–567.

21. Bishop CW, Strugnell SA, Csomor P, et al. Obesity: a key consideration in the management of secondary hyperparathyroidism. Am J Nephrol 2022 (submitted; pending acceptance)

22. Hengist A, Perkin O, Gonzelez JT, et al. Mobilizing vitamin D from adipose tissue: the potential impact of exercise. Nutr Bull 2019; doi: 111/nbu.12369.

23. Comozzi V, Frigo AC, Zaninotto M, et al. 25-Hyderoxycholecalciferol response to single oral cholecalciferol loading in the normal weight, overweight and obese. Osteoporosis Int 2016;27:2593–2602.

24. Michaud J, Naud J, Ouimet D, et al. Reduced hepatic synthesis of calcidiol in uremia. J Am Soc Nephrol 2010;21:1488–1498.

25. Armas LAG, Hollis BW, Heaney RP. Vitamin D2 is much less effective than vitamin D_3_ in humans. J Clin Endocrinol Metab 2004;89:5387–91.

26. Ekwaru JP, Zwicker JD, Holick MF, et al. The importance of body weight for the dose response relationship of oral vitamin D supplementation and serum 25-hydroxyvitamin D in healthy volunteers. Plos One 2014;e111265.

27. Roizen JD, Long C, Casella A, et al. Obesity decreases hepatic 25-hydroxylase activity causing low serum 25-hydroxyvitamin D. J Bone Miner Res 2019;34:1068–1073.

28. Petkovich M, Bishop CW. Extended-Release calcifediol in renal disease. Vitamin D, Volume 2, Health, Disease and Therapeutics, Fourth Edition 2018;667–678.

29. Sitrin MD, Bengoa JM. Intestinal absorption of cholecalciferol and 25-hydroxycholecalciferol in chronic cholestatic liver disease. Am J Clin Nutr 1987;46:1011.

30. Bishop JE, Collins ED, Okamura WH, et al. Profile of ligand specificity of the vitamin D binding protein for 1-alpha,25-dihydroxyvitamin D_3_ and its analogs. J Bone Miner Res 1994;9:1277–88.

31. Didriksen A, Build A Jakobsen, et al. Vitamin D_3_ increase in abdominal subcutaneous fat tissue after supplementation with vitamin D_3_. Eur J Endocrinol 2015;172:235–41.

